# Identification of Novel Genomic Loci for Anxiety and Extensive Genetic Overlap with Psychiatric Disorders

**DOI:** 10.1101/2023.09.01.23294920

**Authors:** Markos Tesfaye, Piotr Jaholkowski, Alexey A. Shadrin, Dennis van der Meer, Guy F.L. Hindley, Børge Holen, Nadine Parker, Pravesh Parekh, Viktoria Birkenæs, Zillur Rahman, Shahram Bahrami, Gleda Kutrolli, Oleksandr Frei, Srdjan Djurovic, Anders M. Dale, Olav B. Smeland, Kevin S. O’Connell, Ole A. Andreassen

**Affiliations:** Centre for Precision Psychiatry, Division of Mental Health and Addiction, Oslo University Hospital, and Institute of Clinical Medicine, University of Oslo, Oslo, Norway; Department of Clinical Science, University of Bergen, Bergen, Norway; KG Jebsen Centre for Neurodevelopmental Disorders, University of Oslo and Oslo University Hospital, Oslo, Norway; Institute of Psychiatry, Psychology and Neuroscience, King’s College London, London, UK; Center for Bioinformatics, Department of Informatics, University of Oslo, Oslo, Norway; Department of Medical Genetics, Oslo University Hospital, Oslo, Norway; Department of Radiology, University of California, San Diego, La Jolla, CA, USA; Multimodal Imaging Laboratory, University of California San Diego, La Jolla, CA, USA; Department of Psychiatry, University of California, San Diego, La Jolla, CA, USA; Department of Neurosciences, University of California San Diego, La Jolla, CA, USA

**Author notes:** **Corresponding authors** Markos Tesfaye, M.D., Ph.D. and Ole Andreassen, M.D., Ph.D. Division of Mental Health and Addiction, Oslo University Hospital & Institute of Clinical Medicine, University of Oslo Building 49, Oslo University Hospital, Ullevål, Kirkeveien 166, PO Box 4956 Nydalen, 0424 Oslo, Norway.

**Keywords:** anxiety, genetic overlap, psychiatric disorder, genetic loci

## Abstract

**Background:** Anxiety disorders are prevalent and anxiety symptoms co-occur with many psychiatric disorders. We aimed to identify genomic risk loci associated with anxiety, characterize its genetic architecture, and genetic overlap with psychiatric disorders.

**Methods:** We used the GWAS of anxiety symptoms, schizophrenia, bipolar disorder, major depression, and attention deficit hyperactivity disorder (ADHD). We employed MiXeR and LAVA to characterize the genetic architecture and genetic overlap between the phenotypes. Conditional and conjunctional false discovery rate analyses were performed to boost the identification of genomic loci associated with anxiety and those shared with psychiatric disorders. Gene annotation and gene set analyses were conducted using OpenTargets and FUMA, respectively.

**Results:** Anxiety was polygenic with 12.9k estimated genetic risk variants and overlapped extensively with psychiatric disorders (4.1-11.4k variants). MiXeR and LAVA revealed predominantly positive genetic correlations between anxiety and psychiatric disorders. We identified 114 novel loci for anxiety by conditioning on the psychiatric disorders. We also identified loci shared between anxiety and major depression (n = 47), bipolar disorder (n = 33), schizophrenia (n = 71), and ADHD (n = 20). Genes annotated to anxiety loci exhibit enrichment for a broader range of biological pathways and differential tissue expression in more diverse tissues than those annotated to the shared loci.

**Conclusions:** Anxiety is a highly polygenic phenotype with extensive genetic overlap with psychiatric disorders. These genetic overlaps enabled the identification of novel loci for anxiety. The shared genetic architecture may underlie the extensive cross-disorder comorbidity of anxiety, and the identified genetic loci implicate molecular pathways that may lead to potential drug targets.

## Introduction

Anxiety is a human emotion while anxiety disorders encompass several categories of mental disorders characterized by the core features of excessive fear and anxiousness, or avoidance behaviors.^1^ Anxiety disorders including generalized anxiety disorder, panic disorder, agoraphobia, social anxiety disorder, and specific phobias are among the leading causes of global disease burden.^2^ Because of the extensive phenotypic overlap and comorbidity among various anxiety disorders,^3^ there is a growing recognition that they are better understood and measured along a continuum rather than discrete categories.^4, 5^ Epidemiological data show frequent comorbidity between anxiety disorders and other psychiatric disorders.^3^ For example, nearly two-thirds of individuals with anxiety disorders also have concurrent depressive disorders.^6^ Anxiety disorders are also common in individuals with bipolar disorder (BIP),^7, 8^ schizophrenia (SCZ),^9^ and attention deficit hyperactivity disorder (ADHD).^10, 11^ Further, symptoms of anxiety (ANX), not meeting the criteria for anxiety disorder, frequently co-occur with other psychiatric disorders.^12, 13^ Anxiety disorders and ANX co-occurring with psychiatric disorders have been linked with greater symptom burden, poorer course and outcome, and lower quality of life.^14–18^ The clinical relevance of co-occurring ANX is highlighted by its inclusion in the diagnostic criteria of ‘with anxious distress’ as a specifier of psychiatric disorders such as major depression (MD), and BIP.^1^

The etiology of anxiety disorders is not clearly understood; however, both genetic and environmental factors are involved.^1, 3, 4, 19, 20^ Genetic susceptibility plays a major role in anxiety disorders,^20^ with heritability estimates from twin studies ranging between 30 to 50%.^21^ Genome-wide association studies (GWAS) have identified several genetic loci for anxiety disorders,^20, 22, 23^ and ANX.^24^ A study has identified 73 loci associated with latent factor of anxiety symptoms.^25^ Recent evidence from genetic correlation analyses supports the notion that there is shared genetic liability between psychiatric disorders, and both anxiety disorders and ANX.^22, 24^ However, genetic correlations do not provide a comprehensive overview of the shared genetic architecture between two phenotypes.^26, 27^ The identification of novel genetic loci for anxiety disorders and a better understanding of the shared genetic landscape with other psychiatric disorders can unveil more information about the biological pathways underlying anxiety disorders.^28, 29^ This is crucial for the development of more effective treatments against anxiety disorders and comorbid conditions. Advances in statistical genetics have improved genetic discoveries for complex diseases.^30, 31^ The utility of these novel analytical methods in characterizing the genetic architecture of anxiety disorders and the genetic overlap with psychiatric disorders may add to our understanding of the underlying biological mechanisms.

The standard approach to evaluate shared heritability in complex disorders is to estimate genetic correlations with linkage disequilibrium score regression (LDSC).^32^ However, genetic correlation, typically a genome-wide summary measure,^31, 33^ may conceal shared genetic architecture involving a mixture of concordant and discordant effect directions and does not capture specific overlapping loci and relevant genes.^26, 27, 34^ These characteristics can be captured by applying bivariate causal mixture model (MiXeR)^26^ and local analysis of covariant association (LAVA).^34^ Genetic overlap can be exploited to boost the discovery of specific genetic risk loci for a phenotype,^31^ and loci shared with other phenotypes by applying conditional false discovery rate (condFDR) and conjunctional FDR (conjFDR) analyses respectively.^30, 35^

Given the high comorbidity between ANX and psychiatric disorders, we aimed to quantify and characterize their genetic overlap. To this end, we applied univariate MiXeR to comprehensively characterize the polygenic architecture of ANX measured using a dimensional scale. We also employed bivariate MiXeR and LAVA to assess the genetic overlap with MD, SCZ, BIP, and ADHD. To identify genetic loci associated with ANX, we performed a meta-analysis of two GWAS and applied the condFDR method. We then performed conjFDR to identify loci shared between ANX and the four psychiatric disorders. Finally, we tested whether polygenic liability to the psychiatric disorders predicted anxiety disorder status in an independent sample.

## Methods and materials

### Genome-wide association for anxiety

We obtained quantitative anxiety trait (ANX) data from the UK Biobank (UKB) measured using the seven-item Generalized Anxiety Disorder (GAD-7) scale in 2016. Genotype data for white British, unrelated participants (n = 126,569) was used for the GWAS. The mean age (SD) of the participants was 64.3 (7.6) years, and 55.9% were female.

### GWAS summary statistics data

We obtained GWAS summary statistics for anxiety symptoms (ANX) measured as a quantitative trait using the Generalized Anxiety Disorder 2-item scale (GAD-2) from the Million Veterans Program (MVP) (*n* = 175,163).^24^ The ANX GWAS summary statistics were accessed through the Database of Genotype and Phenotype (dbGaP; phs001672). We obtained GWAS summary statistics for BIP, ADHD, and SCZ from the Psychiatric Genomics Consortium (PGC),^36–38^ and MD from a meta-analysis of the PGC and 23andMe, Inc.^39^ All the GWAS comprised populations of only European ancestry (Table 1 and *Supplementary Methods* in Supplement 1). All individual studies contributing to these GWAS datasets have been approved by their respective ethical committees.

**Table 1:** GWAS summary statistics data used for investigation of genetic overlap, discovery of genomic risk loci, and polygenic risk prediction.

| Phenotype | Number of cases | Number of controls | Effective sample size | Source of data |
| --- | --- | --- | --- | --- |
| MVP-ANX <sup>a</sup> | - | - | 175,163 | MVP <sup>24</sup> |
| UKB-ANX <sup>a, b</sup> | - | - | 126,569 | UKB |
| META-ANX <sup>a</sup> |  |  | 301,732 | MVP + UKB |
| MD | 246,363 | 561,190 | 684,817 | PGC-MD + 23andMe <sup>39</sup> |
| BIP | 41,917 | 371,549 | 150,670 | PGC-BIP <sup>38</sup> |
| SCZ | 53,386 | 77,258 | 126,282 | PGC-SCZ <sup>37</sup> |
| ADHD | 38,691 | 186,843 | 128,214 | PGC-ADHD <sup>36</sup> |
| ALL ANX <sup>c</sup> | 37,517 | 482,693 | 150,058 | FINNGEN |

GWAS: Genome-wide association study, ANX: Anxiety symptoms, MD: Major depression, BIP: Bipolar disorder, SCZ: Schizophrenia, ADHD: Attention deficit hyperactivity disorder, MVP: Million Veterans Program, PGC: Psychiatric Genomics Consortium, UKB: UK Biobank, META-ANX: meta-analysis of the MVP-ANX and UKB-ANX.
<sup>a</sup> Quantitative trait without case – control dichotomy.
<sup>b</sup> Project No. 27412
<sup>c</sup> Replication dataset comprising GWAS of various anxiety related disorders

### Target sample for polygenic risk score (PRS)

We obtained genotype data for a total of 130,992 population-based cohort of mothers and fathers from the Norwegian Mother, Father, and Child Cohort Study (MoBa).^40^ The MoBa study is conducted by the Norwegian Institute of Public Health and includes approximately 114,500 children, 95,200 mothers and 75,200 fathers. Participants were enrolled from all over Norway from 1999-2008. The details of sample collection, genotyping, and quality control are provided elsewhere.^41, 42^ We obtained ICD-10 psychiatric diagnoses from the Norwegian Patient Registry up until June 2022. Data on a total of 95,841 (58.2 % females) unrelated participants of European Ancestry was used in the analyses. The mean age (SD) of the participants was 49.0(5.5) years. The cases of anxiety disorder (*n* = 4469) comprised agoraphobia (F40.0, *n* = 900), social phobia (F40.1, *n* = 1,345), specific phobia (F40.2 = 432), panic disorder (F41.0, *n* = 1,343), and generalized anxiety disorder (F41.1, *n* = 1,742). MoBa cohort participants had provided written informed consent at enrollment. We excluded participants who withdrew their consent from the analyses. MoBa study was approved by the Regional Committees for Medical and Health Research Ethics (2016/1226).

### Statistical analysis

#### GWAS and Meta-analysis

We performed GWAS of ANX (GAD-7 quantitative scores) among unrelated individuals of European ancestry in the UKB (Project No. 27412). The GWAS was run using PLINK and applied the following filters: minor allele frequency > 0.001, Hardy-Weinberg equilibrium *p*-value > 1.0e-09, and genotyping missingness rate <0.1. Age, sex and the first 20 genotype principal components were included as covariates.

We performed a fixed-effects inverse variance-weighted meta-analysis of the anxiety (ANX) GWAS from the MVP and that from the UKB using METAL.^43^ The resulting summary statistics (*n* = 301,732) were used for the investigation of genetic overlap, identification of ANX loci and shared loci between anxiety (ANX) and psychiatric disorders.

##### Assessing Genetic overlap

We performed a series of MiXeR analyses to investigate the genetic architecture of ANX and its genetic overlap with MD, BIP, SCZ, and ADHD.^26^ First, we conducted univariate MiXeR analyses to estimate the number of trait-influencing variants explaining 90% of SNP-based heritability after controlling for linkage disequilibrium (LD). These were followed by bivariate MiXeR analyses to estimate the number of SNPs shared between pairs of phenotypes irrespective of effect direction. We also determined the estimated proportion of shared SNPs between two phenotypes out of the total number of SNPs estimated to influence both phenotypes (Dice coefficients) and the fraction of SNPs with concordant effects in the shared component.^26^ More detailed information about MiXeR models is provided in the supplement (*Supplementary Methods* in Supplement 1).

We employed LAVA to estimate local genetic correlations between ANX, and MD, BIP, SCZ, and ADHD.^34^ LAVA estimates local genetic correlations and local heritability across 2,495 semi-independent genetic regions of approximately 1Mb and identifies shared genetic regions with their effect directions.^34^ It takes sample overlap into account by using the genetic covariance intercept from LDSC.^32^ LAVA estimates the heritability of each of the genetic regions for each of the phenotypes and then estimates local genetic covariance between pairs of phenotypes.

##### Conditional and conjunctional false discovery rates

We generated quantile-quantile (Q-Q) plots where the *p*-values of single nucleotide polymorphisms (SNPs) in ANX were plotted conditional on three different cut-offs of *p*-values in the secondary phenotypes (i.e., one of MD, BIP, SCZ, and ADHD). Q-Q plots with successive leftward and upward deviation compared to the null were considered to exhibit cross-trait enrichment.^30^ We performed condFDR analyses to identify loci associated with ANX. Next, we applied conjFDR analyses to identify loci shared between ANX and each of the secondary phenotypes respectively.^30, 35^ In condFDR analysis, the SNP *p*-values in the ANX GWAS summary statistics were re-ranked based on their *p*-values in the GWAS summary statistics of the secondary phenotype. CondFDR leverages the SNPs’ association with the secondary phenotype to boost the power to identify novel SNPs associated with the primary phenotype (i.e., anxiety).^30^ This boost in power is contingent on the extent of genetic overlap between the two phenotypes.^30, 44^

We then performed inverse condFDR analyses whereby MD, BIP, SCZ, and ADHD were primary phenotypes, and ANX was the secondary phenotype and used both pairs of condFDR results for conjFDR analyses. The conjFDR value for a SNP is defined by taking the maximum of the condFDR and inverse condFDR values for a given pair of phenotypes.^30^ A threshold of 5% was used as statistically significant for both condFDR and conjFDR *p*-values. We excluded SNPs within the extended major histocompatibility complex (MHC) region and chromosome 8p23.1 inversion (genome build 19 positions of chr6:25119106 – 33854733 and chr8:7200000 – 12500000, respectively) from the condFDR model fit procedure, but not from the discovery analyses.^45^ All the *p*-values were corrected for inflation using a genomic inflation control procedure as described previously (*Supplementary Methods* in Supplement 1).^35^

### Definition of genomic loci

We designated independent genomic loci according to the functional mapping and gene annotation (FUMA) protocol.^46^ We specified candidate SNPs as any SNP with condFDR or conjFDR < 0.05, and candidate SNPs with LD *r*^2^ < 0.6 with each other as independent significant SNPs. Lead SNPs were defined as independent SNPs with LD *r*^2^ < 0.1. The candidate SNPs in LD *r^2^*≥ 0.6 with a lead SNP delineated the boundaries of a genomic locus. We defined all candidate SNPs positioned within the boundaries of a genomic locus to correspond to a single independent genomic locus. We obtained LD information from the 1000 Genomes Project European reference panel.^47^ In conjFDR, we interpreted the effect directions by comparing the *Z*-scores of lead SNPs for each locus in the GWAS summary statistics corresponding to the phenotype. We defined novel risk loci as genomic loci not identified in the GWAS catalog for ANX and anxiety disorder (accessed in January 2024) and in ANX or anxiety disorders GWAS.^20, 23–25, 48–55^

### Consistency of genetic effects in an independent sample

We performed a left-sided binomial test of lead SNPs for concordant effect directions in the discovery (GWAS used for condFDR) and independent dataset from the Finnish population (FinnGen, https://r10.finngen.fi/). The independent dataset comprised GWAS summary statistics of lifetime anxiety disorders based on ICD-10 diagnosis in the Finnish population (Table 1).

### Polygenic risk scores

In MoBa, we restricted the polygenic risk score (PRS) analyses to individuals of European ancestry selected based on genotype principal components (PCs) as described elsewhere.^56^ We used a kinship coefficient greater than 0.05 to exclude one of the related pairs of study participants while prioritizing individuals with anxiety disorders. When two related individuals had an anxiety disorder diagnosis, one of them was selected randomly. We used PRSice^57^ to calculate PRSs at different *p*-value thresholds (i.e., 5e-8, 1e-6, 1e-5, 1e-4, 1e-3, 1e-2, 5e-2, 1e-1, 5e-1, 1) using the GWAS summary statistics for ANX, MD, BIP, ADHD, and SCZ (Table 1). Subsequently, we extracted the first PC for each PRS across all *p*-value thresholds.^58^ Next, we used logistic regression to estimate PRS association with anxiety disorder using models that included PRS of the disorder, and age, sex, and the first 10 genotype PCs as covariates. The combined model also included the PRSs for all the psychiatric disorders and age, sex, and the first 10 genotype PCs as covariates.

### Functional annotations and gene set analyses

We performed functional gene mapping for lead SNPs from cond/conjFDR using the OpenTargets platform (https://genetics.opentargets.org/).^59^ For each lead SNP, we selected the gene with the highest overall score. We used genes mapped to the lead SNPs using the OpenTarget platform for gene set analyses conducted using the GENE2FUNC analyses in FUMA. We also obtained Combined Annotation Dependent Depletion (CADD) scores which show how deleterious the SNP is on protein function,^60^ and RegulomeDB scores which predicted the regulatory function of the SNP from FUMA.^61^ We obtained the expression of genes identified in 54 different human tissues using Genotype-Tissue Expression.^62^

## Results

### Polygenicity and genetic overlap

The MiXeR analyses showed that ANX is a polygenic trait with 12.9k ± 1.2k (mean ± SD) trait-influencing variants contributing to 90% of its heritability (Supplement 2: Table S1). We estimated the SNP heritability of ANX to be 5.1 ± 0.3% (Table 2). The other psychiatric disorders were also polygenic with the following estimated numbers of trait-influencing variants: MD 13.9k ± 0.4k, BIP 8.6k ± 0.2k, SCZ 9.6k ± 0.2k, and ADHD 7.7k ± 0.4k (Supplement 2: Tables S2 – S5), as reported previously.^27, 36^

**Table 2:** SNP heritability and genetic correlation parameters between anxiety and psychiatric disorders from LD score regression analyses.

| Phenotypes | SNP heritability (SE) | Genetic correlation ($r_g$, SE) | $p$-value ($r_g$) |
| --- | --- | --- | --- |
| ANX | 0.051 (0.003) | 1.00 | - |
| MD | 0.065 (0.002) | 0.66 (0.02) | 6.45e-193 |
| BIP | 0.198 (0.008) | 0.24 (0.03) | 4.19e-17 |
| SCZ | 0.384 (0.013) | 0.30 (0.03) | 5.07e-32 |
| ADHD | 0.167 (0.008) | 0.37 (0.03) | 6.32e-33 |

SNP: Single nucleotide polymorphism, LD: Linkage disequilibrium, SE: Standard error, $r_g$ : Genetic correlation, ANX: Anxiety symptoms, MD: Major depression, BIP: Bipolar disorder, SCZ: Schizophrenia, ADHD: Attention deficit hyperactivity disorder.

In the bivariate MiXeR analyses, ANX exhibited a large genetic overlap with psychiatric disorders as demonstrated by the estimated number of shared variants with MD (11.4k ± 1.1k), BIP (8.5k ± 0.3k), SCZ (9.4k ± 0.3k), and ADHD (4.1k ± 0.3k). The Dice coefficients also indicated a substantial overlap with MD (84.7%), BIP (79.3%), SCZ (84.0%), and to a lesser extent with ADHD (39.7%). Most trait-influencing variants shared between ANX, and the psychiatric disorders had concordant effect directions, with 92% for ADHD, 82% for MD, 60% for BIP, and 61% for SCZ (Figure 1, Supplement 2: Tables S6 – S9). Genome-wide genetic correlations using LDSC also showed a significant positive genetic correlation (r) between ANX and all four psychiatric disorders (Table 2).

**Figure 1.**
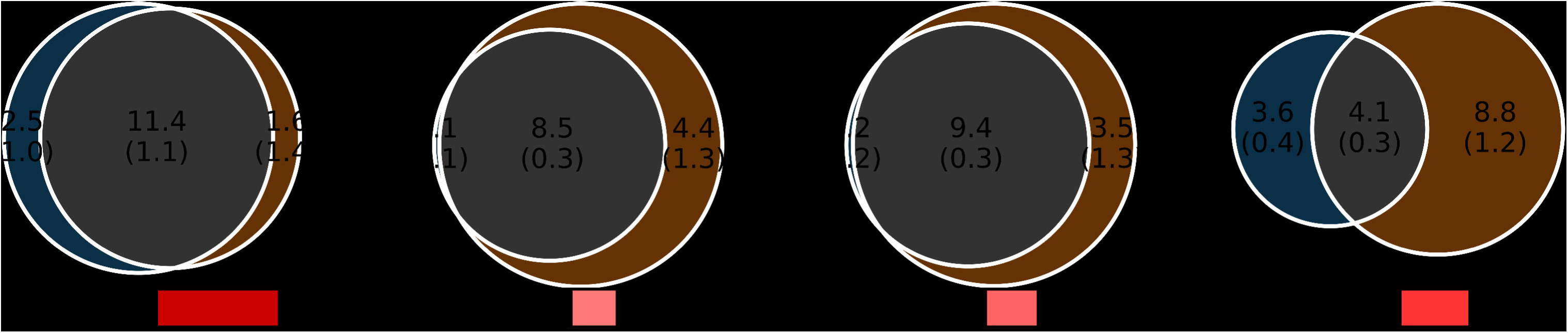
**A – D:** Bivariate MiXeR – Genome-wide genetic overlap between anxiety symptoms (ANX), and major depression (MD), bipolar disorder (BIP), schizophrenia (SCZ), and attention deficit hyperactivity disorder (ADHD). *r_g_*: genetic correlation. The numbers indicate estimates of trait-influencing variants in thousands.

Local genetic correlation estimates from LAVA showed that several regions had positive genetic correlations between ANX and MD (*n* = 17), SCZ (*n* = 6), ADHD (*n* = 3), and BIP (n = 1) after Bonferroni correction. Only one region identified had significant negative genetic correlation between ANX and SCZ (Figure 2; Supplement 1: Table S10).

**Figure 2.**
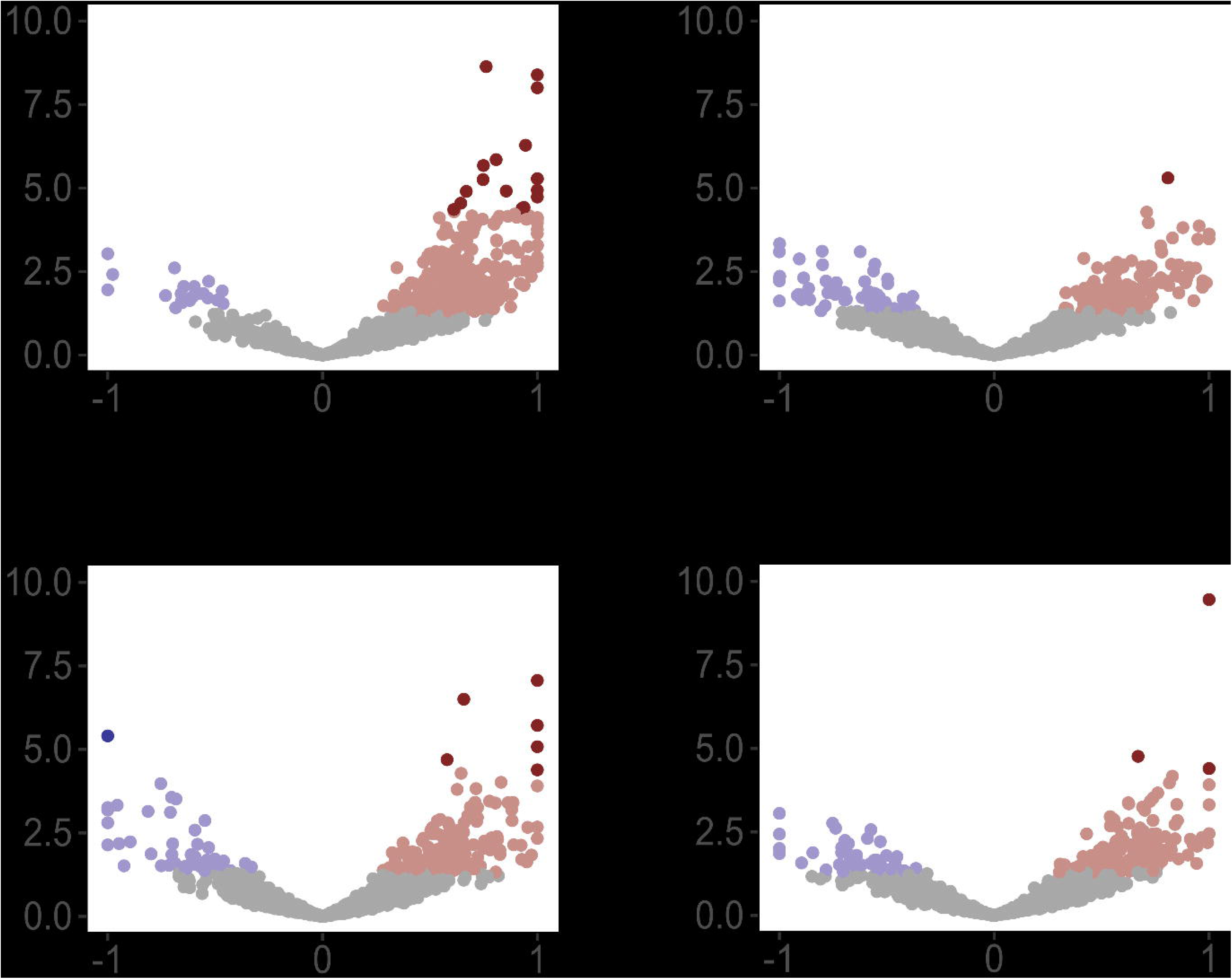
**A – D:** LAVA – Volcano plots of local genetic correlation coefficients (rho) with −log_10_ *p* values for each locus. Dark red dots represent significantly correlated loci after Bonferroni correction. ANX: Anxiety symptoms, ADHD: Attention deficit hyperactivity disorder, BIP: Bipolar disorder, MD: Major depression, SCZ: Schizophrenia.

### Cross-trait polygenic enrichment

We examined the Q-Q plots for cross-trait polygenic enrichment between ANX and psychiatric disorders. The Q-Q plots SNP *p*-values for ANX exhibited upward and leftward deviation when conditioned on progressively smaller *p*-value thresholds from each of MD, BIP, SCZ, and ADHD (Supplement 1: Figure S1). The pattern of Q-Q plots was consistent with the presence of polygenic enrichment between ANX and each of the psychiatric disorders.

### Identification of genetic loci for anxiety

The meta-analysis identified 11 loci associated with ANX of which four were novel (Figure 3, Table 3). Further, we leveraged the cross-trait enrichment and genetic overlap between ANX and psychiatric disorders to identify genomic risk loci for ANX. The condFDR analyses identified 178 unique loci associated with ANX (condFDR < 0.05), 114 of which (64%) were novel (Supplement 3: Tables S11 – S14). The lead SNPs of the identified ANX-loci showed a significant *en masse* concordance of effect directions in the independent GWAS of lifetime anxiety disorders (Table 4).

**Figure 3.**
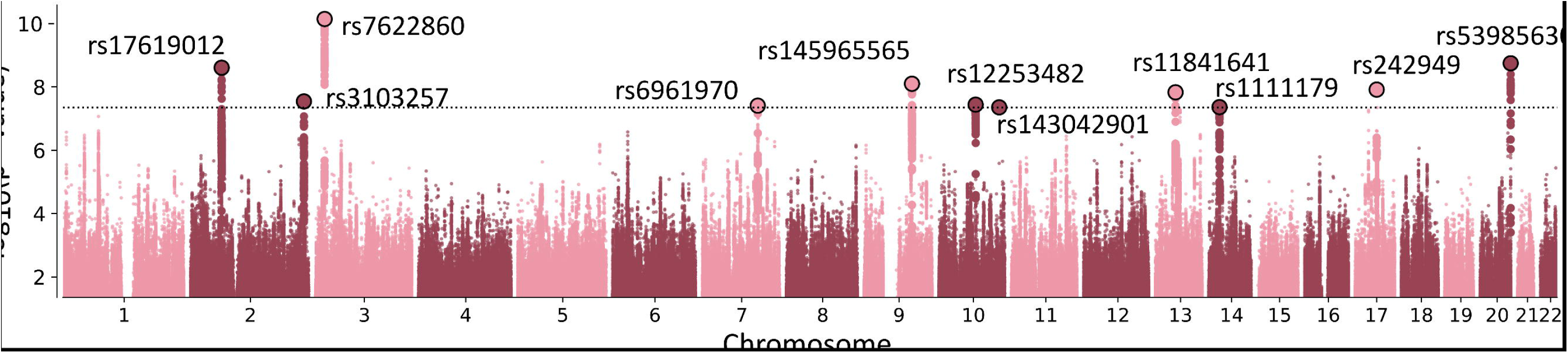
Manhattan plot. Genomic risk loci associated with anxiety. Circled dots indicate the lead single nucleotide polymorphisms with genome-wide significance.

**Table 3:** Genetic risk loci for anxiety identified from meta-analysis of genome-wide association studies.

| CHR: Position<sup>1</sup> | SNP | A1 | A2 | Gene<sup>2</sup> | Function | Beta (SE) | p-value |
| --- | --- | --- | --- | --- | --- | --- | --- |
| 2: 63893589 | rs17619012 | T | G | *WDPCP* | intergenic | -0.023 (0.0039) | 2.80E-09 |
| 2: 233005775 | rs3103257 | A | G | *NPPC* | intronic | 0.020 (0.0036) | 3.23E-08 |
| 3: 18795765 | rs7622860 | A | C | *SATB1* | ncRNA_intronic | 0.023 (0.0036) | 8.09E-11 |
| 7: 113901132 | rs6961970 | A | C | *FOXP2* | intronic | 0.021 (0.0038) | 4.40E-08 |
| 9: 98273305 | rs145965565 | T | G | *PTCH1* | intronic | -0.032 (0.0056) | 8.99E-09 |
| 10: 75496130 | rs12253482 | A | G | *CAMK2G* | ncRNA_intronic | 0.018 (0.0033) | 4.09E-08 |
| 10: 124012277 | rs143042901 | A | G | *PLEKHA1* | intronic | -0.050 (0.0092) | 4.98E-08 |
| 13: 60362163 | rs11841641 | T | C | *DIAPH3* | intronic | 0.040 (0.007) | 1.67E-08 |
| 14: 42075952 | rs1111179 | T | G | *LRFN5* | upstream | -0.018 (0.0032) | 4.90E-08 |
| 17: 43911716 | rs242949 | T | G | *MAPT* | intronic | -0.019 (0.0033) | 1.37E-08 |
| 20: 62714171 | rs539856305 | T | G | *OPRL1* | intronic | 0.034 (0.0057) | 2.02E-09 |

<sup>1</sup>Based on genome build hg19.
<sup>2</sup>Genes annotations were based on the highest overall score in OpenTargets.
CHR Chromosome; A1 Effect allele; A2 Other allele; SNP single nucleotide polymorphism; SE Standard error.
The rows with novel loci are shaded in grey.

**Table 4:**
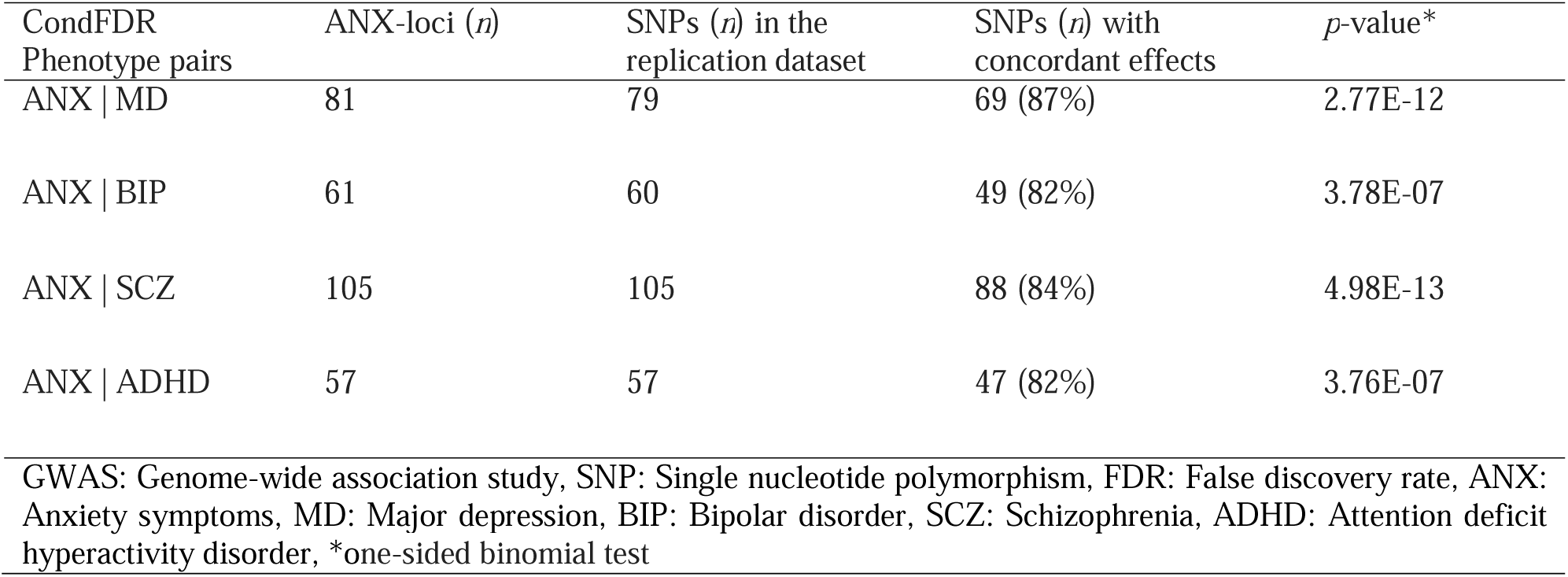
*En masse* test of concordance of effect directions between genetic variants identified for ANX (condFDR) and corresponding variants in independent GWAS of anxiety disorders from the Finnish population (FINNGEN).

### Genomic loci shared between anxiety and psychiatric disorders

We identified 115 genomic loci shared between ANX and psychiatric disorders. Notably, ANX and MD had 47 jointly associated loci (conjFDR < 0.05) all having concordant effect direction. There were 71 loci shared between ANX and SCZ with 52 having concordant effect. Twenty-four of the 33 shared loci between ANX and BIP, and 19 of the 20 loci shared between ANX and ADHD had concordant effects (Supplement 3: Tables S15 – S18). Many of the joint loci were shared across different pairs of traits in conjFDR analysis (Supplement 1: Figure S2).

### Polygenic risk scores

The PRS for each of the psychiatric disorders and ANX were positively associated with anxiety disorders Bonferroni corrected *p*-value < 5.1e-16, MD PRS had a larger effect estimate than all other PRSs. Nagelkerke’s R^2^ showed that MD PRS explained the largest proportion of liability for anxiety disorders (1.52%) followed by ANX PRS (0.41%) (Figure 4A-B; Supplement 4: Table S19). In a multiple regression model where all five PRS were included, the estimates for the associations remained significant for all except PRS of BIP (Supplement 4: Table S20).

**Figure 4.**
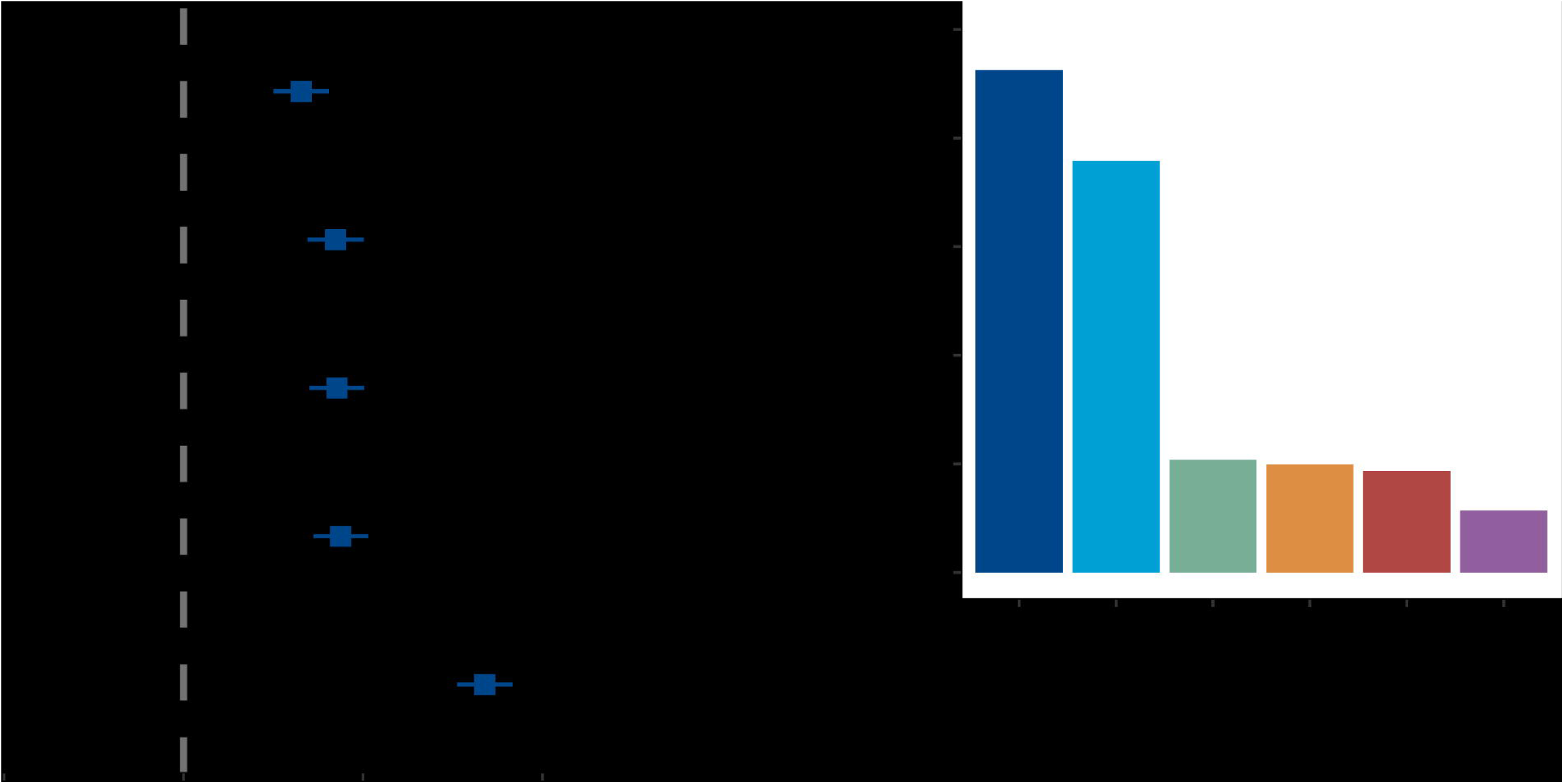
**A – B:** Logistic regression – The association between polygenic risk scores (PRS) of various psychiatric disorders and anxiety disorders in MoBa parents. **A** – models for the PRS of anxiety symptoms (ANX), attention deficit hyperactivity disorder (ADHD), bipolar disorder (BIP), major depression (MD), and covariates. **B** – Nagelkerke’s *R^2^* shows the difference in the percentage of prediction of anxiety traits by each PRS over a base model that includes age, sex, and genotype principal components.

### Functional annotations and gene set analyses

The lead SNPs for the four novel ANX-loci identified in the meta-analysis were intronic and included rs3103257 (*NPPC*), rs6961970 (*FOXP2*), rs143042901 (*PLEKHA1*), and rs11841641 (*DIAPH3*) (Table 3). Two lead SNPs, rs3103257 (*NPPC*) and rs6961970 (*FOXP2*), may have functional significance as indicated by high CADD (Supplement 3: Table S21). Most of the lead SNPs in the ANX-loci identified from condFDR were either intronic or intergenic while four of the lead SNPs were exonic: rs3825393 (*MYO1H*), rs4969391 (*BAIAP2*), rs1468291 (*SERGEF*), and rs61753077 (*TACC2*) (Supplement 3: Tables S11 – S14). Similarly, most lead SNPs in the shared loci were intronic or intergenic except for a SNP shared between ANX and MD - rs3825393 (*MYO1H*), which was exonic. Lead SNPs shared between ANX and MD (rs3793577 and rs3825393), ANX and BIP (rs10497655, rs4702 and rs34961470), ANX and SCZ (rs10497655, rs4702, rs564, rs898031 and rs13262595), and ANX and ADHD (rs61687445 and rs56403421) had a CADD score >12.37 suggesting potential detrimental effects (Supplement 3: Tables S15 – S18).

Gene set analyses of genes annotated to loci identified for ANX with condFDR revealed enrichment of biological processes relevant to neurodevelopment such as neurogenesis (Supplement 5: Table S22). The top enriched cellular components for genes annotated to loci identified for ANX as well as those shared with psychiatric disorders converged to synapse, synaptic membrane, and site of polarized growth (Supplement 5: Tables S23 and S24).

Enrichment analysis of the genes annotated to ANX loci showed that they are differentially expressed in the brain, renal cortex, adrenal gland, vascular, gastrointestinal, and adipose tissues. In contrast, enrichment analysis of the genes annotated to the loci shared between ANX and the psychiatric disorders showed differential tissue expression in the brain and cardiovascular tissues, and renal cortex (Supplement 1: Figures S3 - S4).

## Discussion

Here, we showed that anxiety symptoms are highly polygenic with nearly thirteen thousand trait-influencing genetic variants, with extensive genetic overlap with other psychiatric disorders beyond their positive genetic correlations. Overlapping trait-influencing variants were the largest for ANX and MD, BIP, and SCZ, and a relatively smaller overlap between ANX and ADHD. The proportion of variants with concordant effects within the shared component was the highest for ADHD (92%) and lowest for BIP (60%). Local genetic correlations revealed predominantly positively correlated regions. We identified 114 novel genetic loci associated with ANX, and 115 unique genetic loci shared between ANX and psychiatric disorders, with a similar pattern of effect directions, and a predominance of concordant effects. Consistent with these, polygenic liabilities for the different psychiatric disorders predicted a lifetime clinical diagnosis of anxiety disorders in an independent population-based sample lending further evidence for a shared genetic risk.

We found that ANX is a highly polygenic phenotype. This is important as common genetic variants contribute to a large portion of its heritability.^20^ We also found that common genetic variants in ANX exhibit low discoverability and hence require very large sample size studies for genome-wide discoveries. This may partly explain the observed difference between the SNP heritability we found or reported in other GWASs,^20, 24^ and the PRS performance. Our comprehensive characterization of the genetic overlap between ANX, and MD, BIP, SCZ, and ADHD disorders using methods agnostic to effect directions such as bivariate MiXeR expand our understanding beyond genetic correlations.^24^ Genetic correlations alone are high for ANX and other internalizing disorders^24^ whereas our results from MiXeR revealed extensive overlap with externalizing disorders as well. Further, the higher genetic correlation between ANX and ADHD despite a smaller genetic overlap compared to that of ANX and BIP is probably accounted for by the similar effect directions for most shared genetic variants with ADHD.^26^ The local genetic correlations are consistent with those reported previously.^63^ However, we found larger numbers of correlated genomic regions between ANX, and MD, SCZ and ADHD probably due to the larger power of GWAS data used in the current study.

By leveraging genetic overlap with psychiatric disorders, we obtained more than 16-fold boost in the identification of genetic loci for ANX. Researchers have previously reported a boost in discovery of loci for ANX by leveraging other traits (e.g., neuroticism) using a different approach.^25^ The identification of novel loci revealed biological pathways potentially involved in the pathophysiology of ANX.^46^ Genes annotated to the ANX-loci were enriched for pathways linked to neurodevelopment and cellular components related the synapse. These highlight the role of neurodevelopmental factors in the pathophysiology of anxiety.^64^ Furthermore, the pathways related to the synaptic structures are relevant to the identification of potential drug targets.^65^ Similarly, both the large number of shared genetic loci identified between ANX, and psychiatric disorders and the enriched biological pathways suggest shared mechanisms related to neurotransmission. We speculate that such shared biological pathways may underlie the higher prevalence of anxiety among individuals with psychiatric disorders than in the general population.

The identification of novel loci offers valuable insight into the biology of ANX and their potential molecular mechanisms, especially in relation to comorbid conditions. For example, the gene *FOXP2* encodes a transcription factor that plays a crucial role in regulating gene expression in the human brain.^66^ Mutations in the gene have been associated with speech-language disorder,^67^ and are also linked to a heightened risk of anxiety and depressive disorders.^68^ Similarly, *NPPC,* which encodes a preproprotein for natriuretic peptides, may contribute to the association between anxiety disorders and cardiovascular diseases.^69^ Previous research has shown that natriuretic peptides can alleviate panic attacks,^70^ suggesting a potential therapeutic avenue for anxiety disorders.^71, 72^ Additionally, a study has found an inverse correlation between plasma levels of atrial natriuretic pro-peptide and anxiety in patients with severe heart failure.^73^ The protein coding gene *DIAPH3*, involved in cell adhesion and motility, and is known to play a critical role in cortical neurogenesis,^74, 75^ further highlighting its relevance to mental disorders. Furthermore, *PLEKHA1* has been implicated in both depressive symptoms and type 2 diabetes indicating pleiotropy.^76^ Lastly, *MYO1H,* identified in GWAS of anxiety disorders among individuals of European ancestry,^55^ has also been associated with hereditary spastic paraplegia.^77^ Overall, these findings align with observations that anxiety disorders frequently coexist with various psychiatric and somatic conditions. Further, they provide novel insight into the underlying shared molecular pathways and may assist in improving the treatment of not only comorbid anxiety disorders but also for optimizing treatment for concomitant symptoms of anxiety.

Notably, the genes annotated to ANX loci showed differential tissue expression in a much broader range of tissues including the brain, gastrointestinal, cardiovascular, and endocrine tissues while those of the shared loci. While these may be due to comorbidity between anxiety disorders and medical conditions,^78, 79^ we argue that ANX has stronger somatic component involving various organ systems than other psychiatric disorders. Also, the genetic risk for ANX may influence risk through a more diverse set of tissues than other psychiatric disorders as demonstrated in animal models.^80^

The measures of ANX i.e., the GAD-7 and its shorter version GAD-2 are well established tools with comparable psychometric properties.^81^ ANX as a dimensional trait in the GWAS may have the advantage of capturing several of the anxiety disorders as well as subsyndromal ANX^81^ both of which have important clinical implications. The recognition of concomitant symptoms of anxiety is highlighted by the addition of a diagnostic specifier - anxious distress - in DSM-5.^1, 82,83^ Anxious distress can have a negative impact on the severity and clinical outcome of primary psychiatric disorders.^82, 84^

We acknowledge that the primary phenotype ANX GWAS^24^ was a dimensional trait defined based on self-reported GAD-2 or GAD-7 and referred to symptoms experienced in the preceding two weeks rather than a lifetime clinical diagnosis of specific anxiety disorders. This may have contributed to the low proportion of polygenic liability for lifetime anxiety disorders explained by the ANX PRS. Since individuals with a history of diagnosis of depression were not excluded from the ANX GWAS, the genetic overlap between ANX and MD could partly be due to comorbidity. We applied our analyses to GWAS data from populations of European ancestry and therefore, generalizations cannot be made to other ancestries. As multi-ancestry GWAS data become available, our methods can then be applied to improve genetic discoveries for anxiety.

In conclusion, our investigation of the genetic architecture of symptoms of anxiety revealed a high polygenicity and low discoverability. There was also a large genetic overlap between anxiety and psychiatric disorders, which enabled the identification of 114 novel anxiety loci and 115 shared loci. The shared genetic architecture may underlie the high burden of anxiety symptoms in individuals with other psychiatric disorders. The genetic risk for anxiety may involve pathophysiology in neurodevelopment and neurotransmission. The genes annotated to anxiety loci implicated a broader range of biological pathways as well as differential tissue expression in more diverse tissues than the shared loci. Our findings advance our understanding of the pathophysiology of anxiety that occurs alone or concomitantly with other psychiatric disorders and may help in the identification of potential drug targets. Further research is needed to investigate the genetic underpinnings of specific types of anxiety disorders.

## Data Availability

GWAS of anxiety can be accessed at https://www.ncbi.nlm.nih.gov/gap/, dbGaP Study Accession phs001672, All PGC data are available at https://www.med.unc.edu/pgc/download-results/, Full GWAS summary statistics for the 23andMe DEP dataset will be available through 23andMe to qualified researchers under an agreement with 23andMe that protects the privacy of the 23andMe participants. Interested investigators should and reference this paper for more information. Access to the MoBa data can be obtained by applying to the Norwegian Institute of Public Health (NIPH). Restrictions apply regarding the availability of the MoBa data, and therefore, it is not publicly available. Access can be given after approval provided that the applications are consistent with the consent provided by participants. Detailed information on the application can be found on the NIPH website at https://www.fhi.no/en/studies/moba/. The cond/conjFDR code is available online at https://github.com/precimed/pleiofdr.

https://www.med.unc.edu/pgc/download-results/

https://www.fhi.no/en/studies/moba/

https://github.com/precimed/pleiofdr/

## List of acronyms

ADHD: Attention Deficit Hyperactivity Disorder,
ANX: Anxiety symptoms,
BIP: Bipolar Disorder,
CADD: Combined Annotation Dependent Depletion,
CondFDR: Conditional FDR,
ConjFDR: Conjunctional FDR,
dbGAP: Database of Genotype and Phenotype,
FDR: False Discovery Rate,
FUMA: Functional Mapping and Annotations,
GAD-2: Generalized Anxiety Disorder 2-item scale,
GAD-7: Generalized Anxiety Disorder 7-item scale,
GWAS: Genome-Wide Association Study,
LD: Linkage Disequilibrium,
LDSC: Linkage Disequilibrium Score Regression,
MiXeR: bivariate causal mixture model,
MD: Major Depression,
MHC: major histocompatibility complex,
MVP: Million Veterans Program,
MoBa: Mor og Barn (Norwegian Mothers and Children Cohort),
PGC: Psychiatric Genomics Consortium,
Q-Q: Quantile-Quantile,
SCZ: Schizophrenia,
SNP: Single Nucleotide Polymorphism,
UKB: United Kingdom Biobank

## Declarations

### Ethics approval and consent to participate

The MoBa cohort has initially been approved by the Norwegian Data Protection Agency and The Regional Committees for Medical and Health Research Ethics in Norway and is currently regulated by the Norwegian Health Registry Act. The use of MoBa data for this work was approved under (REK 2016/1226). Individual studies comprising the published datasets have been approved by their respective ethical approval committees. This research was conducted according to the Helsinki Declaration.

### Consent for publication

Not Applicable.

### Availability of data and materials

GWAS of anxiety can be accessed at https://www.ncbi.nlm.nih.gov/gap/, dbGaP Study Accession phs001672, All PGC data are available at https://www.med.unc.edu/pgc/download-results/, Full GWAS summary statistics for the 23andMe DEP dataset will be available through 23andMe to qualified researchers under an agreement with 23andMe that protects the privacy of the 23andMe participants. Interested investigators should email and reference this paper for more information. Access to the MoBa data can be obtained by applying to the Norwegian Institute of Public Health (NIPH). Restrictions apply regarding the availability of the MoBa data, and therefore, it is not publicly available. Access can be given after approval provided that the applications are consistent with the consent provided by participants. Detailed information on the application can be found on the NIPH website at https://www.fhi.no/en/studies/moba/. The cond/conjFDR and MiXeR codes are freely available online at https://github.com/precimed/pleiofdr and https://github.com/precimed/mixer, respectively.

### Competing interests

Ole A. Andreassen is a consultant for Cortechs.ai and Precision Health, and has received speaker’s honoraria from Lundbeck, Janssen, Otsuka and Sunovion. Srdjan Djurovic has received speaker’s honoraria from Lundbeck. Anders M. Dale was a Founder of and holds equity in CorTechs Labs, Inc, and serves on its Scientific Advisory Board. He is also a member of the Scientific Advisory Board of Human Longevity, Inc. (HLI), and the Mohn Medical Imaging and Visualization Centre in Bergen, Norway. He receives funding through a research agreement with General Electric Healthcare (GEHC). The terms of these arrangements have been reviewed and approved by the University of California, San Diego in accordance with its conflict-of-interest policies. The other authors have no conflicts of interest to declare.

### Funding

We gratefully acknowledge support from the American National Institutes of Health (NS057198, EB00790), the Research Council of Norway (RCN) (296030, 324252, 324499, 300309, 273291, 223273), the South-East Norway Regional Health Authority (2022-073), The University of Oslo, and KG Jebsen Stiftelsen (SKGJ-MED-021). This project has received funding from the European Union’s Horizon 2020 research and innovation program under grant agreements No 847776, 964874, and 801133 under the Marie Skłodowska-Curie grant. The MoBa study is research is part of the HARVEST collaboration, supported by the Research Council of Norway (#229624). We also thank the NORMENT Centre for providing genotype data, funded by the Research Council of Norway (#223273), South East Norway Health Authorities and Stiftelsen Kristian Gerhard Jebsen. We further thank the Center for Diabetes Research, the University of Bergen for providing genotype data and performing quality control and imputation of the data funded by the ERC AdG project SELECTionPREDISPOSED, Stiftelsen Kristian Gerhard Jebsen, Trond Mohn Foundation, the Research Council of Norway, the Novo Nordisk Foundation, the University of Bergen, and the Western Norway Health Authorities

### Author contributions

M.T., K.S.O., and O.A.A. conceived and designed the analysis. K.S.O., Z.R., O.F., A.M.D., and O.A.A. contributed to analysis tools. M.T., P.J., K.S.O., D.V.M., and A.S. performed the analyses. M.T. wrote the first draft of the manuscript. All authors contributed to the interpretation of the findings, provided critical intellectual content, and approved the final manuscript.

## Acknowledgments

The Norwegian Mother, Father and Child Cohort Study is supported by the Norwegian Ministry of Health and Care Services and the Ministry of Education and Research. We thank the Norwegian Institute of Public Health (NIPH) for generating high-quality genomic data. We are grateful to all the participating families in Norway who take part in this on-going cohort study. We would like to thank the research participants and employees of 23andMe, the Psychiatric Genomic Consortium (Schizophrenia, Bipolar Disorder, and Depression Working Groups), the UK Biobank, the Million Veteran Program, and MoBa for making this work possible. This work was performed on the TSD (Tjeneste for Sensitive Data) facilities, owned by the University of Oslo, operated and developed by the TSD service group at the University of Oslo, IT-Department (USIT).

The article has been posted on a preprint server - https://doi.org/10.1101/2023.09.01.23294920

## Notes

### Competing Interest Statement

Ole A. Andreassen is a consultant for Cortechs.ai and Precision Health and has received speaker honoraria from Lundbeck, Janssen, Otsuka and Sunovion. Srdjan Djurovic has received speakers honoraria from Lundbeck. Anders M. Dale was a Founder of and holds equity in CorTechs Labs, Inc, and serves on its Scientific Advisory Board. He is also a member of the Scientific Advisory Board of Human Longevity, Inc. (HLI), and the Mohn Medical Imaging and Visualization Centre in Bergen, Norway. He receives funding through a research agreement with General Electric Healthcare (GEHC). The terms of these arrangements have been reviewed and approved by the University of California, San Diego in accordance with its conflict-of-interest policies. The other authors have no conflicts of interest to declare.

### Summary of Updates

The GWAS of the primary phenotype i.e. anxiety now included a large sample from the UKB based on GAD-7 in addition to the MVP GWAS sumstats. Accordingly, all results including MiXeR, CondFDR, ConjFDR, LAVA, LDSC, and Polygenic Risk Scores have been revised using the new summary statistics of anxiety GWAS meta-analysis. The novelty check for genetic risk loci of anxiety has been updated.

