## Supplementary File 1 for "Identification of Novel Genomic Loci for Anxiety and Extensive Genetic Overlap with Psychiatric Disorders"

### Affiliations

### * Corresponding authors

### **Supplementary Methods**

#### **Summary statistics**

##### Dataset for Anxiety Symptoms (ANX)

The GWAS summary statistics for anxiety symptoms (ANX) were obtained from the Million Veteran Project (MVP) cohort. A detailed description of the MVP cohort characteristics is available elsewhere ^1^. In the GWAS of ANX, the phenotype was assessed using a dimensional self-report survey using the GAD-2 scale in 175,163 adults of European Ancestry ^2^. The GAD-2 asks respondents how often they have been bothered by: *Feeling nervous*, *anxious or on edge*, and *not being able to stop or control worrying during the preceding two weeks*. The responses are rated from zero – ‘*not at all*’ to three – ‘*nearly every day*’. The total scores on GAD-2 range from zero to six ^3^. Several studies have reported that GAD-2 had acceptable psychometric properties in the detection of various anxiety disorders ^3, 4^. Overall, none of the study participants had a history of schizophrenia or bipolar disorder. However, individuals with self-reported depression diagnoses (22%) and attention deficit hyperactivity disorder diagnoses (2.3%) were not excluded.

##### Dataset for Major Depression

The GWAS summary statistics for major depression (MD) comprised a meta-analysis of three GWASs of depressive disorders in populations of European Ancestry ^5^. Cases of depression were ascertained using structural clinical interviews or similar criteria for the Psychiatric Genomics Consortium (PGC) (43,204 cases and 95,680 controls) ^6^, the GWAS of self-reported history of diagnosis of depression from 23andMe, Inc. (75,607 cases and 231,747 controls) ^7^, and a broadly defined depression phenotype from the UK Biobank (127,552 cases and 233,763 controls) ^8^.

##### Dataset for Bipolar Disorder

The GWAS summary statistics for bipolar disorder (BIP) comprise 57 cohorts collected in Europe, North America, and Australia from the third wave of the PGC bipolar disorder. The GWAS included 41,917 cases defined as individuals meeting one of the international consensus criteria (DSM-IV, ICD-9, or ICD-10) for a lifetime diagnosis of BIP based on structured diagnostic instruments and 371,549 controls of European Ancestry ^9^. The BIP case status of some cohorts obtained from biobanks was ascertained using ICD codes or self-reports.

##### Dataset for Schizophrenia

The GWAS summary statistics for schizophrenia (SCZ) comprised the European subset of the PGC meta-analysis of cohorts of schizophrenia and schizoaffective disorder. The sample used for this GWAS included 53,386 cases and 77,258 controls ^10^.

##### Datasets for Attention Deficit Hyperactivity Disorder

The GWAS summary statistics of attention deficit hyperactivity disorder (ADHD) included 38,691 cases and 186,843 controls of European Ancestry ^11^. The GWAS was a meta-analysis of GWASs of ADHD including the PGC-ADHD, iPSYCH, and deCODE. The PGC-ADHD included 11 cohorts from different countries and with various ascertainment criteria for ADHD, and in total comprised 4,515 cases and 11,702 controls. The iPSYCH ADHD cases (*n* = 25,895) were obtained from the Danish Psychiatric Central Research Register and were ascertained by psychiatrists to have met the ICD10 criteria for F90.0, F90.1, or F98.8 while controls (n=37,148) were a random sample of individuals from the nationwide birth cohort who have not received a diagnosis of ADHD. The deCODE GWAS defined cases of ADHD as those who met ICD 10 criteria (F90, F90.1, F98.8) (n=5,583), or those who received specific medications for symptoms of ADHD (n=2,698). Individuals in the control group did not have a diagnosis of SCZ, BIP or autism spectrum disorders.

#### **Statistical Analyses**

#### MiXeR Analyses

##### Univariate MiXeR

For each SNP, $i$, MiXeR models its additive genetic effect of allele substitution,$\beta_{I}$, as a point-normal mixture, $\beta_{i}=\left( 1-\pi_{1} \right)N\left( 0,0 \right)+\pi_{1}N\left( 0, \sigma_{\beta}^{2} \right)$, where $\pi_{1}$ represents the proportion of non-null SNPs (i.e., polygenicity) and $\sigma_{\beta}^{2}$ represents variance of effect sizes of non-null SNPs (i.e., discoverability). Then, for each SNP, $j$, MiXeR incorporates LD information and allele frequencies for input SNPs calculated based on 1000 Genomes Phase3 reference panel ^12^ to estimate the expected probability distribution of the signed test statistic, $z_{j}=\delta_{j}+\epsilon_{j}=\sqrt{N}\sum_{i} \sqrt{H_{i}}r_{ij}\beta_{i}+\epsilon_{j}$, where $N$ is sample size, $H_{I}$ indicates heterozygosity of *i*-th SNP, $r_{ij}$ indicates allelic correlation between *i*-th and *j*-th SNPs, and $\epsilon_{j}\mathcal{\sim N}\left( 0, \sigma_{0}^{2} \right)$ is the residual variance. Further, the three parameters, $\pi_{1}, \sigma_{\beta}^{2}, \sigma_{0}^{2}$, are fitted by direct maximization of the likelihood function. The number of trait-influencing variants is estimated as $M\pi_{1}$, where $M$ is the number of SNPs in the reference panel. The phenotypic variance explained on average by a trait-influencing variant is calculated as $\bar{H}\sigma_{\beta}^{2},$ where $\bar{H}=\frac{1}{M}\sum_{i} H_{i}=0.2075$ is the average heterozygosity across SNPs in the reference panel. Under the assumptions of the MiXeR model, SNP-heritability is then calculated as $h_{SNP}^{2}=M\pi_{1}\times\bar{H}\sigma_{\beta}^{2}$ ^13^.

##### Bivariate MiXeR

MiXeR models additive genetic effects as a mixture of four components, representing null SNPs in both traits ($\pi_{0})$; SNPs with a specific effect on the first and on the second trait ($\pi_{1}$ and $\pi_{2}$, respectively); and SNPs with non-zero effect on both traits ($\pi_{12}$). In the last component, MiXeR models variance-covariance matrix as $\boldsymbol{\Sigma}_{\boldsymbol{12}}=\left[ \begin{matrix} \sigma_{1}^{2} & {\rho_{12}\sigma}_{1}\sigma_{2} \\ {\rho_{12}\sigma}_{1}\sigma_{2} & \sigma_{2}^{2} \end{matrix} \right]$ where $\rho_{12}$ indicates correlation of effect sizes within the shared component, and $\sigma_{1}^{2}$ and $\sigma_{2}^{2}$ correspond to the discoverability parameters estimated in the univariate analyses of the two traits. After fitting parameters of the model genetic correlation is calculated as $r_{g}=\frac{\rho_{12}\pi_{12}}{\sqrt{\left( \pi_{1}+\pi_{12} \right)\left( \pi_{2}+\pi_{12} \right)}}.$

The Dice coefficient (DC) was calculated using the formula DC *=* $\frac{2\pi_{12}}{{\pi_{1}+\pi_{2}+2\pi}_{12}}$.

MiXeR performed 20 iterations of each univariate and bivariate analysis with 2 million randomly selected SNPs with a MAF threshold of 5% followed by random pruning at an LD threshold of *r^2^* = 0.8, resulting in a sample of ~600K input SNPs per iteration. Those input SNPs were used to construct the distribution of genetic effects used to calculate the observed signed test statistics. The mean and standard deviation estimates for each parameter were calculated from the sample of 20 iterations ^13^.

To identify analyses using insufficiently powered GWAS summary statistics, MiXeR computes the Akaike information criterion ($AIC=2k-2\ln\left( L \right)$), where $k$ is the number of free parameters in the model and $L$ is the value of the likelihood function. In univariate analysis, the ΔAIC represents the difference between AIC of the model fitted using MiXeR and AIC of the infinitesimal model, which assumes that all variants are non-null resulting in the constrained univariate model with 2 free parameters ($\sigma_{\beta}^{2}, \sigma_{0}^{2}$) while fixing polygenicity parameter equal to 1. A positive ΔAIC value indicates the input GWAS dataset provides sufficient power to discriminate the model fitted using MiXeR from the infinitesimal model.

In bivariate analysis, we calculated the difference between $\mathrm{AIC}$ between the full bivariate model, $k=3$, and the reduced bivariate model, $k=2$, due to $\pi_{12}$ being constrained to the smallest or largest possible value ( $\pi_{12}^{min}=r_{g}\sqrt{\pi_{1}^{u} \pi_{2}^{u}}$ and $\pi_{12}^{max}=\min\left( \pi_{1}^{u}, \pi_{2}^{u} \right)$*),* respectively. Each bivariate analysis returns two ΔAIC values, including ΔAIC_best_vs_min_ (which compares MiXeR modelled fit with a constrained model with minimal polygenic overlap) and ΔAIC_best_vs_max_ (which compares MiXeR modelled fit with a constrained model with maximum polygenic overlap). When both ΔAIC_best_vs_min_ and ΔAIC_best_vs_max_ are positive, it implies that the GWAS summary statistics have enough information to distinguish the modelled polygenic overlap versus the constrained models with minimal ($\pi_{12}^{min}$) and maximum ($\pi_{12}^{max}$) polygenic overlap. A positive ΔAIC_best_vs_min_ with a negative ΔAIC_best_vs_max_, which were observed for ANX and BIP or ANX and SCZ (Supplementary Table 1), imply genetic overlap beyond the minimal polygenic overlap and suggest further investigations using more powerful GWAS datasets will help finalize the estimates of polygenic overlap ^13^.

##### Conditional and Conjunctional False Discovery Rates (CondFDR/ConjFDR)

###### Conditional Q-Q plots

Q-Q plots of nominal p-values from GWAS summary statistics are one of the common approaches through which one can visualize the enrichment of statistical association relative to that expected under the global null hypothesis (none of genetic variants influences the phenotype). The Q-Q curve has the nominal p-values, denoted by “p”, on the y-coordinate and the corresponding value of the empirical cumulative distribution function (cdf), denoted by “q”, on the x-coordinate. The -log_10_(p) is commonly plotted against the -log_10_(q) to accentuate tail probabilities of the theoretical and empirical distributions. Under the global null hypothesis, the theoretical distribution of p-values is uniform on the interval [0,1]. In the presence of all null relationships, nominal –log10(p) form a straight (diagonal) line on a Q-Q plot when plotted against the empirical distribution of –log10(q). Leftward deflections of the observed distribution from the projected null line reflect increased tail probabilities in the distribution of test statistics and consequently an over-abundance of low p-values compared to that expected by chance, termed as ‘enrichment’.

Conditional Q-Q plots are constructed by creating subsets of SNPs based on levels of an auxiliary measure for each SNP, and computing Q-Q plots separately for each cut-off of the auxiliary measure ^14^. In our analysis, the significance of association with the secondary (conditional) phenotype is used as an auxiliary measure. If SNP enrichment is obtained because of variation in the auxiliary measure, this is expressed as successive leftward deflections in a conditional Q-Q plot as levels of the auxiliary measure increase. The enrichment can be directly interpreted in terms of the true discovery rate (1−FDR) ^15^. Cross-trait enrichment exists if the proportion of SNPs associated with a phenotype increases as a function of the strength of the association with a secondary phenotype.

We constructed conditional Q-Q plots of empirical quantiles of nominal -log10 p-values for SNP association for all SNPs, and for subsets (strata) of SNPs determined by the nominal p-values of their association with the conditional phenotypes, and vice versa. Specifically, we computed the empirical cumulative distribution of nominal p-values for ANX for all SNPs and for SNPs with significance levels below the indicated cut-offs for the conditional phenotypes (e.g., SCZ) at p < 0.1, p < 0.01, p < 0.001. The nominal p-values (– log10(p)) are plotted on the y-axis, and the empirical cumulative distribution function of nominal p-values (–log10(q), where q=ecdf(p)) is plotted on the x-axis.

###### Conditional and Conjunctional FDR

CondFDR is an extension of the standard FDR, which incorporates information from GWAS summary statistics of a second (conditional) phenotype to adjust significance levels in the first (primary) phenotype. The condFDR is defined as the probability that a SNP is null in the first phenotype given that the p-values in the first and second phenotypes are as small as or smaller than the observed ones. The condFDR estimates are obtained for each nominal SNP p-value in the primary phenotype after computing the stratified empirical cdfs of the nominal p-values ^16, 17^. The separate strata are determined by the relative enrichment of SNP associations as a function of increased nominal SNP p-values in a secondary phenotype. The standard FDR framework derives from a model that assumes that the distribution of test statistics in a GWAS can be formulated as a mixture of null and non-null effects, with true associations having more extreme test statistics than false associations on average. Ranking SNPs by the standard FDR or by p-values gives the same ordering of SNPs. Given the cross-trait SNP enrichment between the primary and secondary phenotypes, the condFDR procedure re-orders SNPs and results in a different ranking than that using p-values alone.

ConjFDR analysis aims to boost the statistical power for the discovery of shared genetic variants between two phenotypes. It is defined as the posterior probability that a SNP is null for either phenotype or both simultaneously, given that its p-values for association with both phenotypes are as small as or smaller than the observed p-values ^18, 19^. We implement conjFDR analysis as two consequent condFDR analyses. First, ANX is the primary phenotype and one of the others (e.g., SCZ) is the conditional (secondary) phenotype. Second, an inverse condFDR is performed where ANX becomes the conditional phenotype and the other phenotype (e.g., SCZ) becomes the primary phenotype. Then a conservative estimate of the conjFDR values for each SNP can be derived as the maximum of two condFDR values ^14^. The significance threshold for the conjFDR analysis was defined as a conjFDR value < 0.05. More methodological details of condFDR and conjFDR are available in the original publications and a more recent review ^14, 18^.

### **Supplementary Figures**

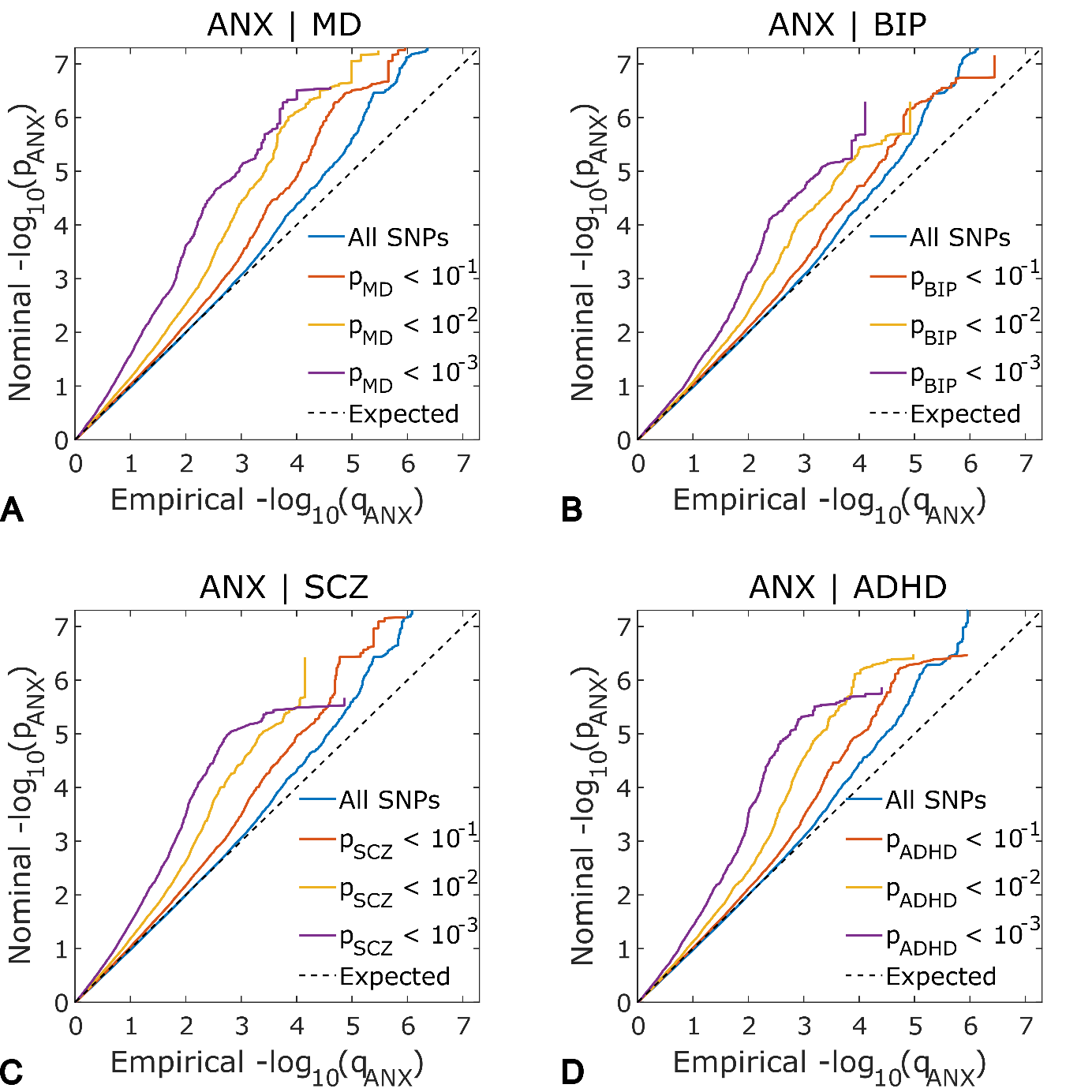

**Figure S1.** Conditional Q-Q plots of nominal *-log10 p*-values vs empirical *-log10 p*-values for SNP associations with anxiety symptoms (ANX) conditional on the *p*-values for their association with major depression (MD), bipolar disorder (BIP), schizophrenia (SCZ) and attention deficit hyperactivity disorder (ADHD).

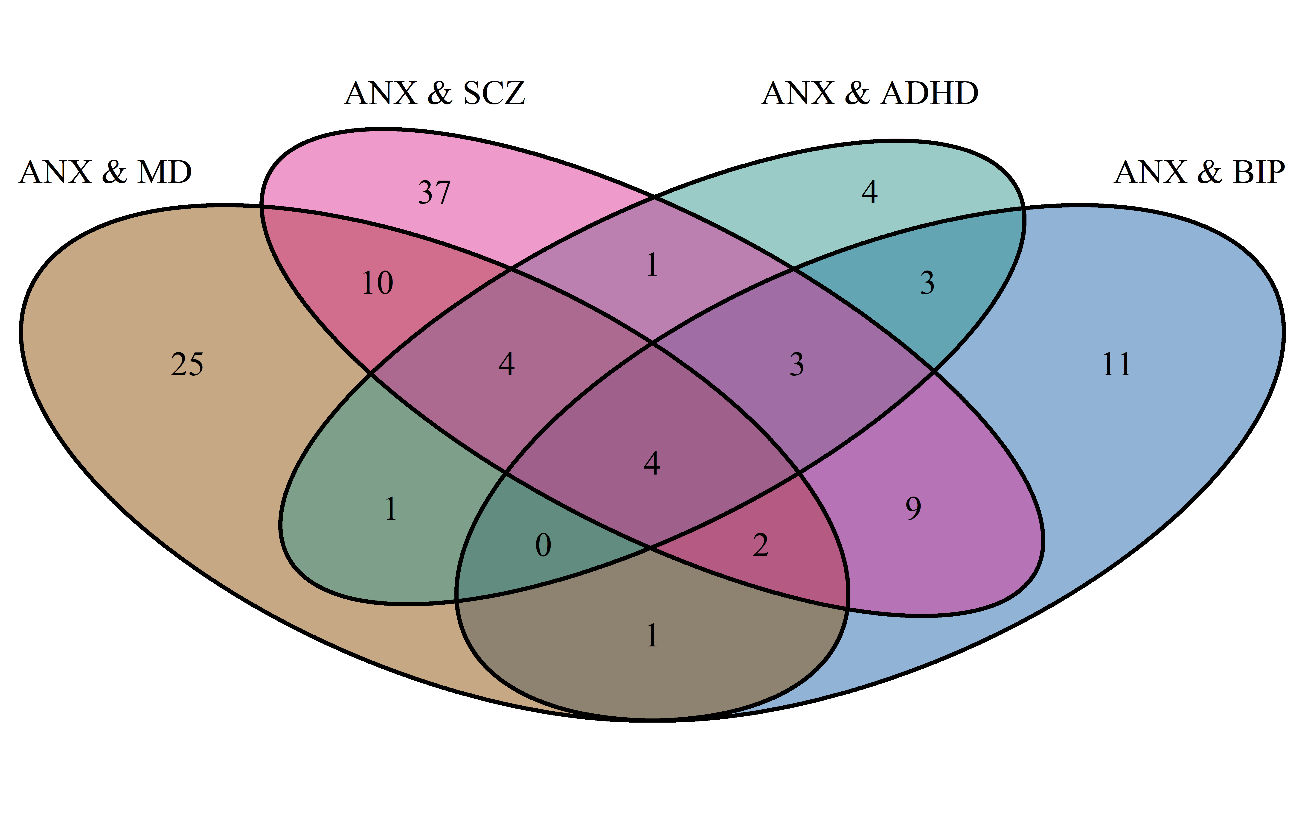

**Figure S2.** Overlap among shared loci identified for anxiety symptoms (ANX) conjunctional with psychiatric disorders: Major depression (MD), bipolar disorder (BIP), schizophrenia (SCZ), and attention deficit hyperactivity disorder (ADHD). Numbers indicated the numbers of shared loci using conjFDR method.

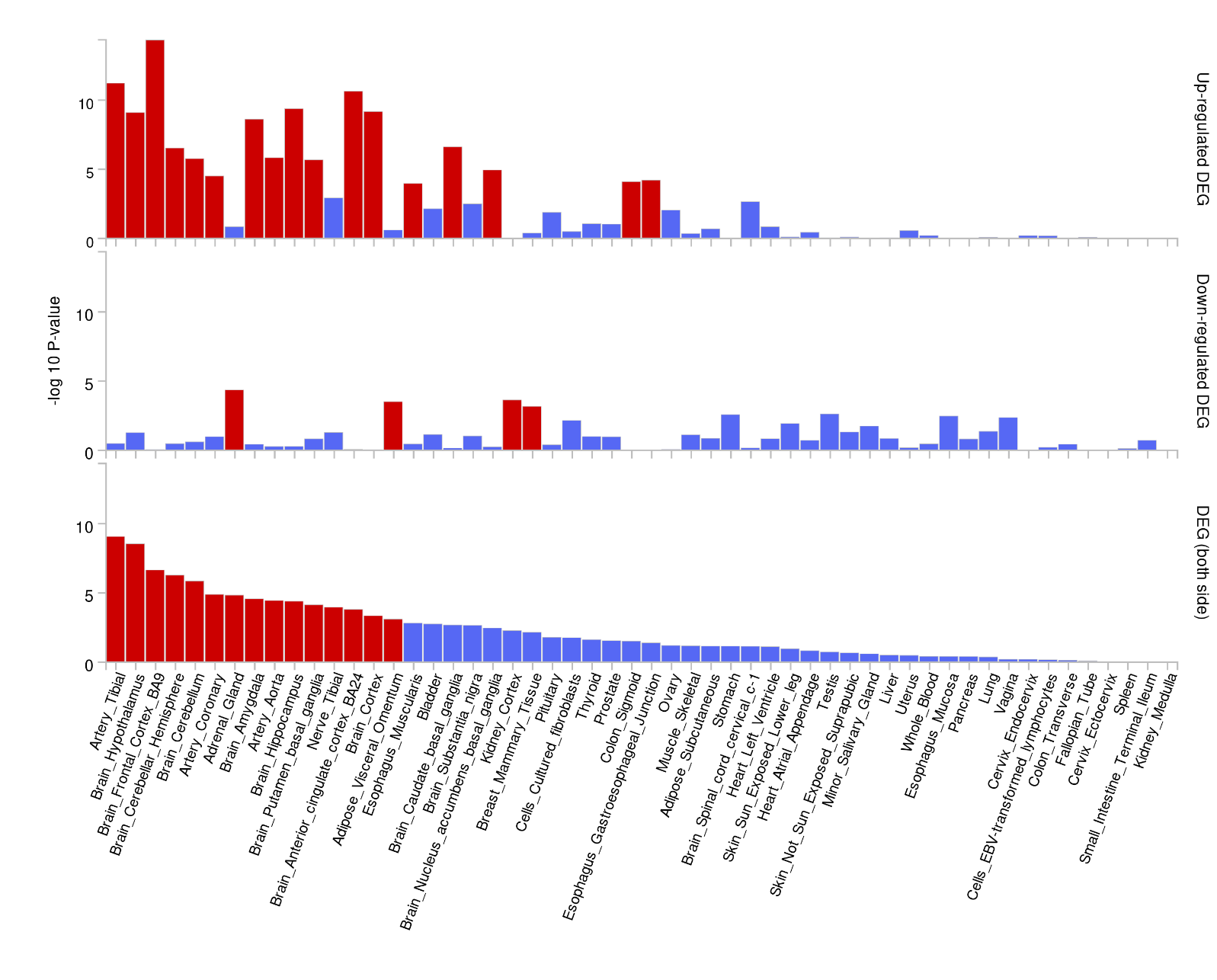

**Figure S3.** FUMA functional annotation – Tissue enrichment for differential gene expression (DEG) in 54 GTEx tissue types of genes mapped to genomic loci associated with anxiety symptoms (condFDR < 0.05; Supplement 3).

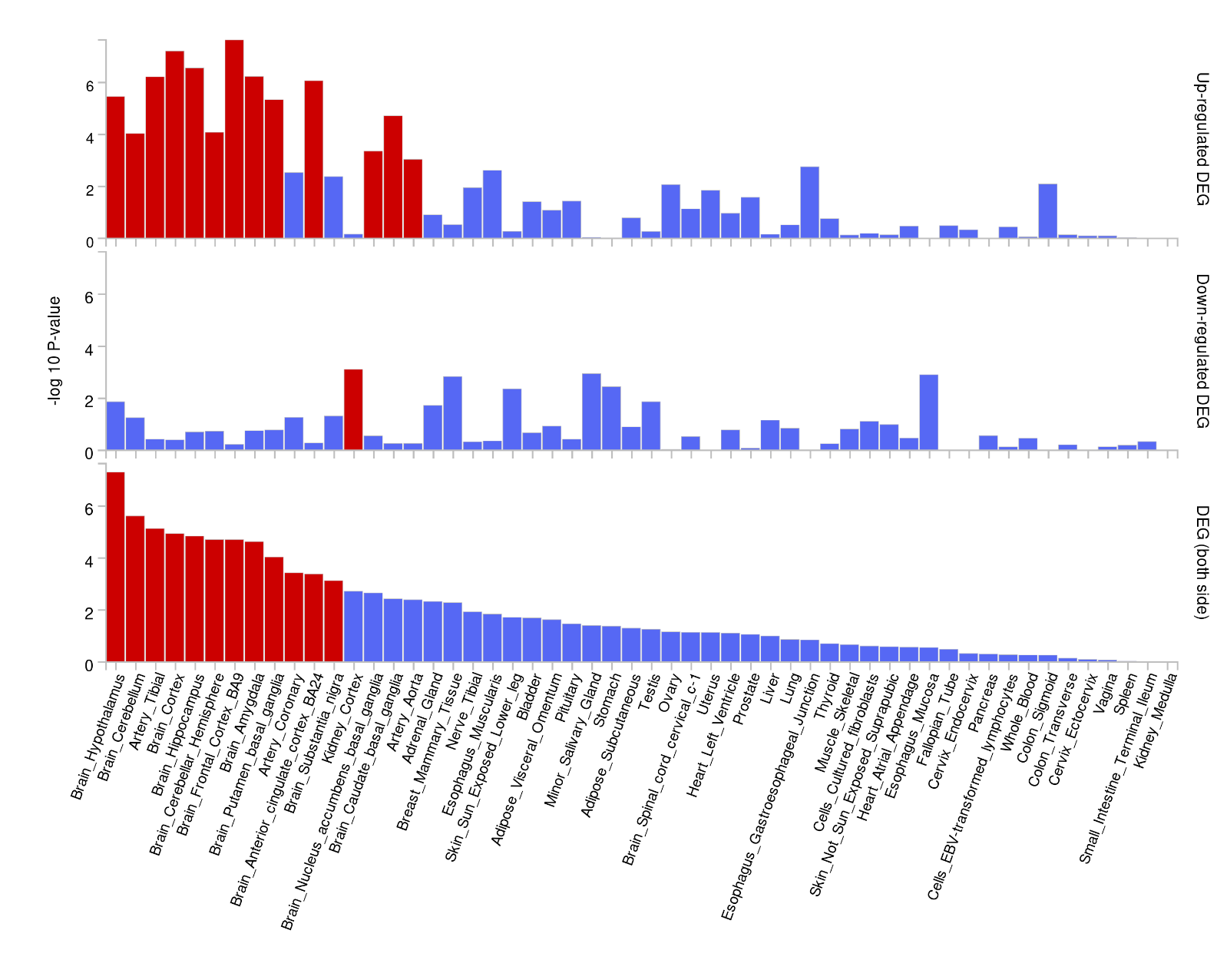
**Figure S4.** Tissue enrichment for differential gene expression (DEG) in 54 GTEx tissue types of genes mapped to genomic loci jointly associated with anxiety, and psychiatric disorders (major depressive disorder, bipolar disorder, schizophrenia, and attention deficit hyperactivity disorder) (conjFDR< 0.05; Supplement 3).

| **Table S10.** LAVA – Number of positively and negatively correlated genomic regions between anxiety symptoms and attention deficit hyperactivity disorder (ADHD), bipolar disorder (BIP), major depression (MD), and schizophrenia (SCZ). | | | | | |
| --- | --- | --- | --- | --- | --- |
| **Genomic regions in the univariate LAVA** | | **Genomic regions in the Bivariate LAVA** | | | |
| **Anxiety** | **Secondary Phenotype** | **Total** | **Significant**  **(**P_bonferroni_ <0.05**)** | **Concordant** | **Discordant** |
| 2119 | MDD: 2367 | 1109 | 17 | 17 | 0 |
| 2129 | BIP: 2364 | 1109 | 1 | 1 | 0 |
| 2135 | SCZ: 2398 | 1179 | 7 | 6 | 1 |
| 2067 | ADHD: 2315 | 952 | 3 | 3 | 0 |
